# HIV-1 founder variant multiplicity is determined by the infection stage of the source partner

**DOI:** 10.1101/19013524

**Authors:** Ch. Julián Villabona-Arenas, Matthew Hall, Katrina A. Lythgoe, Stephen G. Gaffney, Roland R. Regoes, Stéphane Hué, Katherine E. Atkins

## Abstract

During sexual transmission, the large genetic diversity of HIV-1 within an individual is frequently reduced to one founder variant that initiates infection ^1^. Understanding the drivers of this bottleneck is crucial to develop effective infection control strategies ^2^. Genetic characteristics of the potential founder viruses and events in the recipient partner are both known to contribute to this bottleneck, but little is understood about the importance of the source partner ^3^. To test the hypothesis that the source partner affects the multiplicity of HIV founder variants, we developed a phylodynamic model calibrated using genetic and epidemiological data on all existing transmission pairs for whom the direction of transmission and the infection stage of the source partner are known. Our results demonstrate the importance of infection stage of the source partner, and not exposure route, in determining founder variant multiplicity. Specifically, acquiring infection from someone in the acute (early) stage of infection increases the risk of multiple variant transmission when compared with someone in the chronic (later) stage of infection. This study provides the first direct test of source partner characteristics to explain the low frequency of multiple founder strain infections and can inform clinical intervention study design and interpretation.

## Main

Sexual transmission of HIV-1 results in a viral diversity bottleneck due to physiological barriers as well as viral or cellular constraints that prevent most genetic variants within the source partner from establishing onward infection ^1,4,5^. Indeed, this diversity bottleneck results in around three quarters of new infections being founded by a single genetic variant ^2,6–10^. The extent of genetic diversity transmitted to a new partner is a crucial determinant in understanding the efficacy of putative vaccines and may shed light on the transmission of drug resistance to treatment naive individuals.

The factors leading to the diversity bottleneck during sexual transmission can be broadly categorized as those determined by the source partner—such as viral load and viral diversity available for transmission, those determined by the recipient partner—such as target cell type and availability in the genital or rectal mucosa (e.g. ^3,5,11^), and those connected with viral characteristics—such as glycosylation profiles and cell tropism (reviewed in ^12^). While the impact of the recipient partner and the characteristics of transmitted variants have been widely discussed, little is known about how the source partner affects the viral diversity bottleneck. In particular, despite the importance of infection stage as a driver of HIV transmission—that is, the length of time between the source partner becoming infected and transmission to their partner—there is no empirical evidence to suggest how this influences the viral diversity bottleneck. This gap has arisen because analyses are routinely conducted on individuals without information on the partner from whom they acquired infection. Phylogenetic analyses now offer a possible solution to this impasse.

Phylogenetic trees are representations of the ancestral relationships of organisms with the tips of the tree representing those that are sampled, the internal nodes their inferred common ancestors, and the branches as the evolutionary pathways between these actual and inferred individuals. When phylogenetic trees are constructed using sequence data from both partners in an HIV transmission pair, the relationship between the evolutionary histories of both set of viral samples may reflect epidemiological relationships between the two individuals ^13–15^. Previous modelling studies suggest that by assigning the relationship between the evolutionary histories of both partners as one of three topology classes—monophyletic-monophyletic (MM), paraphyletic-monophyletic (PM), or a combination of paraphyletic and polyphyletic (PP)— epidemiological information can be inferred, such as the direction of transmission ^13^, as well as evolutionary information, such as determining the number of transmitted variants ^16^. That is, the number of monophyletic clusters in a PM (one) or PP (more than one) tree can be interpreted as the minimum number of transmitted lineages (**Fig. 1A**). In practice, however, many factors may influence epidemiological interpretations from phylogenetic trees such as sampling times, sampling density of the viral populations and phylogenetic signal ^17,18^.

**FIGURE 1:**
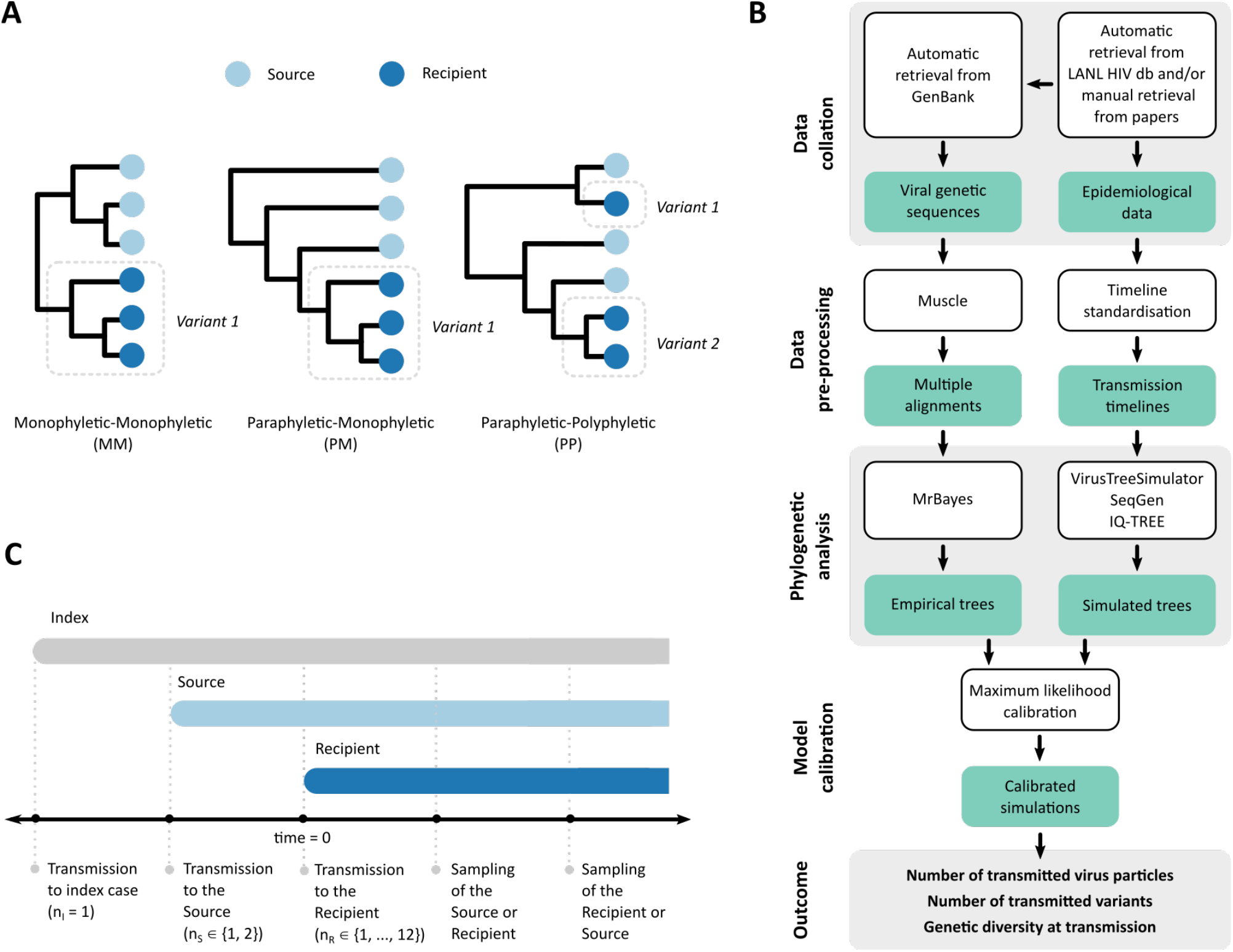
Methods schematics. A) Phylogenetic tree topology class of known transmission pairs that have previously been used as a proxy for calculating the minimum number of founder variants transmitted to the recipient: trees of class MM and PM both suggest a minimum of one founder variant while trees of class PP suggest a multiple founder variants, with the minimum number of founder variants being the number of recipient clades embedded in PP trees (here shown as two). B) Pipeline of phylodynamic analysis (LANLdb, Los Alamos National Laboratory HIV sequence database) where teal represents data or analysis output and white represents methods and analysis. An example of a standardised transmission timeline for a known source-recipient pair is provided in panel C. C) Schematic of the transmission pair model simulation that shows the transmission and sampling timelines. The simulated number of virus particles transmitted to the index case, and the source and recipient partners (*n*_I_, *n*_S_, *n*_R_ respectively) are shown on the transmission events timeline.

Here we present a data-driven phylodynamic approach to overcome these empirical and methodological issues to evaluate the impact of the source partner’s infection stage and route of exposure on the HIV diversity bottleneck (**Fig. 1B**,**C**). We first retrieved all available genetic and epidemiological information from published HIV sexual transmission pairs where the direction of transmission is known, and kept for further analysis those pairs for whom transmission could be classified as having occurred in the source partner’s acute stage (≤90 days after his/her infection) or chronic stage (later than 90 days after his/her infection). After further stratifying pairs into heterosexual (HET) and men-who-have-sex-with-men (MSM) risk groups, we found a significant difference in the timing of transmission between the two risk groups. Specifically, 10 of 36 MSM pairs were the result of acute stage transmission compared with 1 of 76 of HET pairs (**Fig. 2**).

**FIGURE 2:**
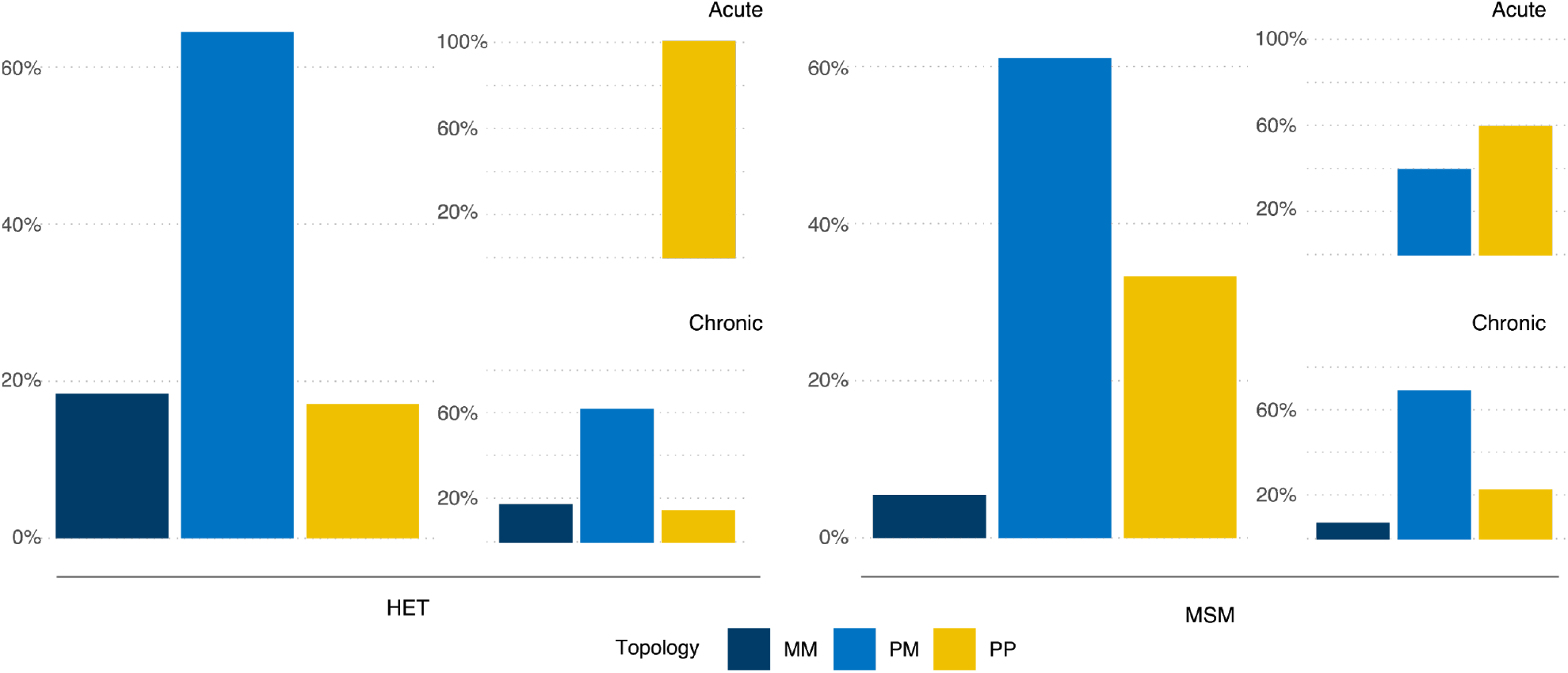
Phylogenetic findings from the empirical transmission pairs. Fraction of phylogenetic tree topology class (MM: Monophyletic-Monophyletic, PM: Paraphyletic-Monophyletic and PP: Paraphyletic-Polyphyletic) where each tree topology class is classified as the most frequent topology class of each posterior distribution per transmission pair. Results are stratified by risk group: 76 heterosexual (HET) pairs and 36 men-who-have-sex-with-men (MSM) pairs) and infection stage of the source partner at transmission (11 acute pairs defined as <90d post infection and 101 chronic pairs defined as ≥90d post infection).

We then performed Bayesian phylogenetic tree reconstruction on the genetic sequences of the transmission pairs and classified the topology class of each tree in the posterior distribution as monophyletic-monophyletic (MM), paraphyletic-monophyletic (PM) or paraphyletic-polyphyletic (PP). The most likely topology class was PM (65% and 61% for HET and MSM, respectively), but with a higher number of PP trees in the MSM group (*P*=0.056, **Fig. 2**). This result has previously been reported as indicative of a higher number of founder variants for MSM ^16^. However, when we stratify the topology class by whether the source partner was in acute or chronic infection at the time of transmission, our results indicate that the infection stage of the source is the primary driver for any observed differences in topology class. Specifically, there is no difference between the HET and MSM groups in the PM/PP topology class ratio when transmission occurs in the chronic stage of infection (*P*=0.570). Note that only one HET transmission occurs during the acute stage, and the topology class for this pair is PP. These results remain qualitatively consistent when only data were analysed from the 66% of transmission pairs for whom the posterior trees gave a certainty of over 95% for the most frequent topology class (**Fig. S3**). These results indicate that infection stage of the source partner, and not risk group *per se*, influences the diversity bottleneck at transmission.

To test whether these empirical findings are indicative of a smaller diversity bottleneck in the chronic stage of HIV infection, we developed a phylodynamic framework in which we simulated the epidemiologic characteristics of each HET and MSM transmission pair, the timing of their sequence sampling, the transmission of virus particles, and the within-host genetic evolution in both the source and recipient (**Fig. 1B)**. Specifically, using the epidemiological information from the transmission pairs, we simulated phylogenies under a coalescent model before generating genetic sequences from these simulations and performing Maximum Likelihood (ML) phylogenetic reconstruction on these simulated sequences. We classified each of these simulated trees as MM, PM or PP and determined the frequency of each topology class for each simulated transmission pair across all the simulated sequences. However, as we could not directly observe the number of virus particles that are transmitted between source and recipient, we repeated the simulation of phylogenetic trees for each transmission pair under a range of plausible values of virus particles transmitted. By fitting the simulation output topology class distribution to the topology class distribution from the empirical phylogenetic trees using maximum likelihood inference, we then determined the most likely number of transmitted virus particles for each transmission pair and used this best fit model for further analysis. Note that two or more virus particles may have the same genetic sequence, and would constitute a single founder variant (or haplotype), discussed later.

Our fitting procedure selects a best fit model that clearly delineates between transmission pairs between whom one virus particle is transmitted (75% of pairs) and those between whom more than one virus particle is transmitted (25% of pairs, **Fig. 3A**). While there is a high degree of confidence in the result when one particle is transmitted, there is often uncertainty around the exact number when multiple particles are transmitted (**Fig. 3A**). Importantly, we found acute stage transmissions are less likely to lead to single particle infections compared with chronic stage transmissions (27% vs. 80%, *P* = 0.0005). The topology class of the simulated phylogenetic trees is strongly influenced by the number of virus particles being transmitted (**Fig. 3B**). PM trees are more commonly found in the pairs that are better described by a model with a single transmitted virus particle (81%) whereas PP trees appeared more often when multiple particles are likely to have been transmitted (86%).

**FIGURE 3:**
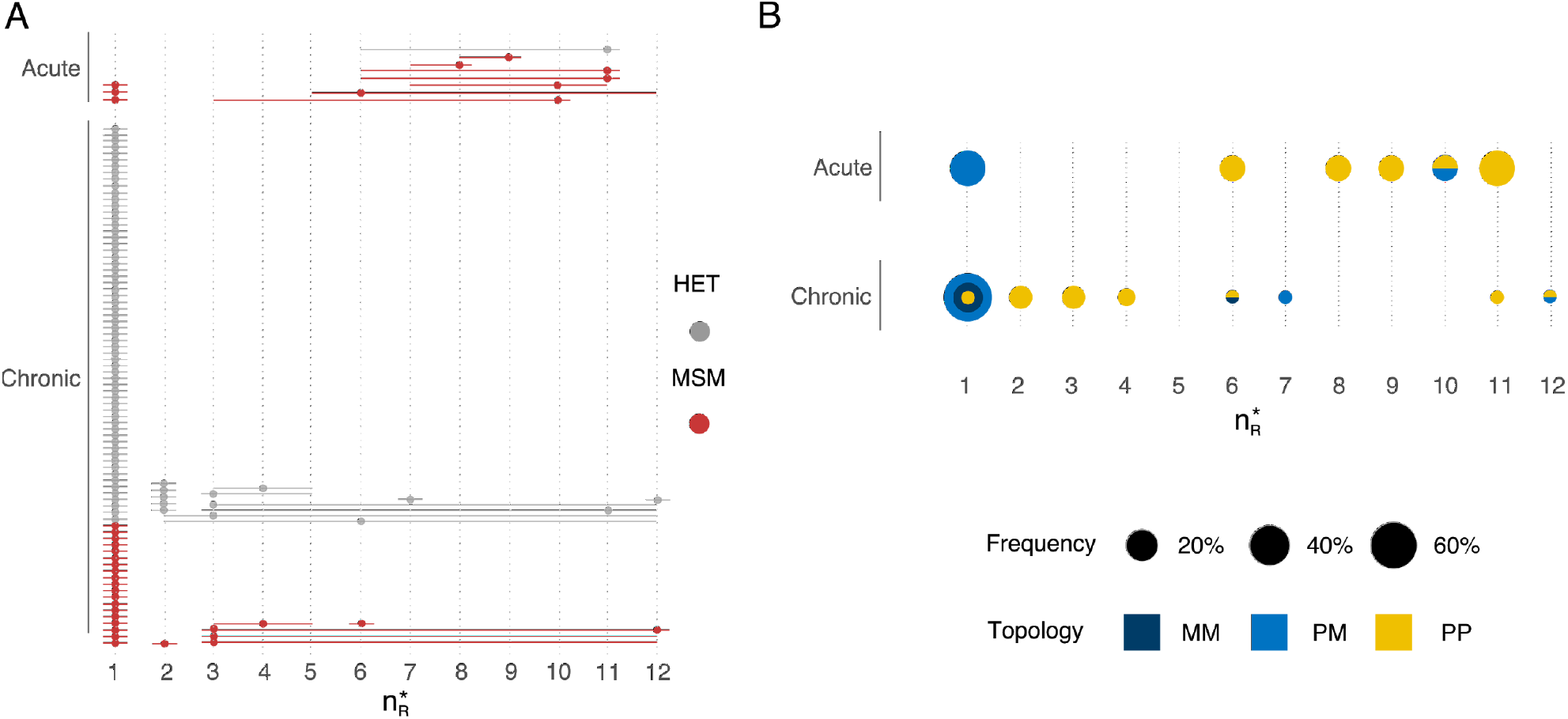
The estimated number of transmitted virus particles for the 112 transmission pairs. The estimates of transmitted virus particles for each transmission pair were calculated by choosing the model simulation that generated a phylogenetic tree topology class distribution (that is, the number of MM, PM and PP trees constructed from the simulated genetic sequences) that best matched the topology class distribution from the phylogenetic trees constructed from the empirical genetic sequences. A) Maximum likelihood number of virus particles founding recipient infections, 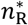, for each pair (stacked points) with 95% confidence intervals (lines) grouped by stage of infection (acute, 11 pairs or chronic, 101 pairs) and risk group (76 heterosexual pairs, HET and 36 men-who-have-sex-with-men pairs, MSM). B) Maximum likelihood number of virus particles founding recipient infections coloured by topology class of the phylogenetic tree constructed from the simulated genetic sequences.

For each transmission pair, we then simulated the genetic sequences of the transmitted viral population under the best fit virus particle model and calculated the most likely number of transmitted variants for each transmission pair. The median number of variants transmitted across all pairs is 1 (range: 1-11, **Fig. 4A**). Using the full distribution of founder variant multiplicity for each pair, we also calculated the probability that a single founder variant was transmitted to the respective recipient. Our results suggest that across all pairs in both risk groups, the mean probability of observing one founder variant is 0.73. Stratifying by risk group, we find there is a higher probability that one variant founds HET infections than MSM infections (a geometric mean of 0.80 vs. 0.63, **Fig. 4B**). However, these risk group differences mostly disappear when we stratify the results by the infection stage of the source. Here, for example, when only chronic stage transmissions are considered, the probability of one founding variant is a little higher for MSM transmissions than for HET transmissions (means of 0.80 vs 0.71), and the pairwise diversity at transmission is similar between both groups (**Fig. 4C**). In contrast, when stratifying solely by infection stage of the source partner, we find that transmission during the acute stage has a much lower probability of one founder variant than during the chronic stage (means of 0.40 vs. 0.77) with a higher median number of variants transmitted, when only the most likely multiplicity for each pair is considered (2 vs. 1, **Fig. 4A**). Nonetheless, if multiple variant transmission does occur, our results suggest that the number of founder variants is higher during chronic stage transmission, consistent with a higher diversity measure during this later stage of infection (**Fig. 4C**).

**FIGURE 4:**
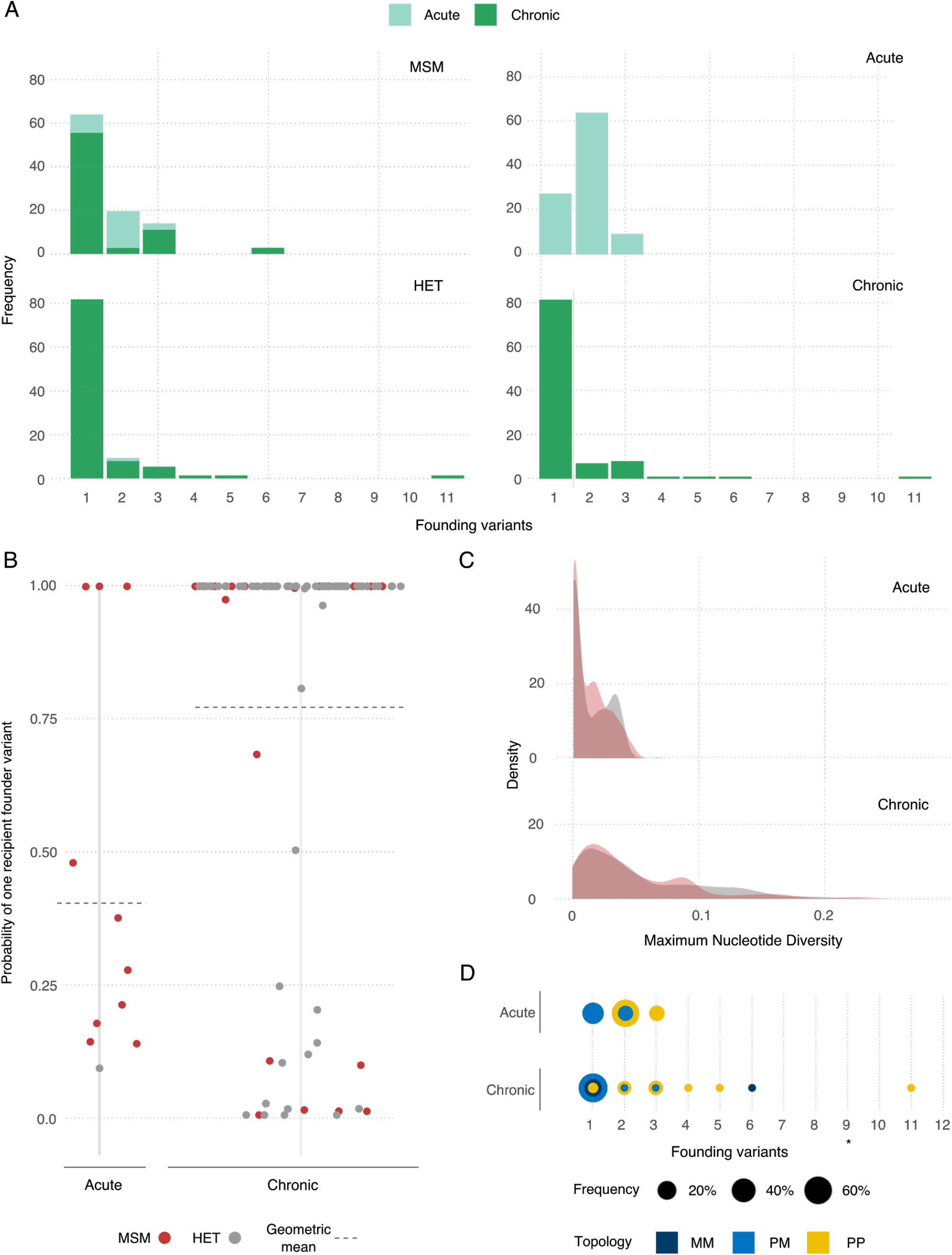
Phylogenetic findings from the calibrated simulations. A) Frequency of multiplicity of founding variants for transmission pairs by infection stage of source partner at transmission and risk group. The multiplicity is calculated as the modal simulated value. B) Probability of one founder variant in the recipient for each pair stratified by infection stage of the source partner at transmission. C) Probability density distribution of maximum diversity (proportion of sites that differ) in the recipient partner across all simulations with more than one haplotype stratified by infection stage of the source at transmission. D) Number of founding variants coloured by topology class of the phylogenetic tree constructed from the best fit model of the simulated genetic sequences.

From these results, therefore, there is approximately double the chance of multiple variant transmission during acute stage infection across both risk groups (relative risk = 0.52). Assuming that transmission risk is weighted towards early transmission such that half of all index case to source partner transmissions occur after 90 days of index case infection leads to qualitatively similar results (Supplementary Information). Similarly, calibrating the simulation model to bootstrapped samples rather than Bayesian posterior distributions leads to similar results (Supplementary Information).

Our results suggest that there is an association between tree topology class and multiple variant transmission, with 95% of MM and PM trees being due to one founder variant (**Fig. 4D**). However, the number of embedded recipient clades is not always a proxy for the minimum number of founder variants transmitted. For example in chronic stage transmission, 11% of PP topology class trees were due to single variant transmission (**Fig. 4D**). Across both infection stages, we find that if MM, PM or PP is assigned as the most likely tree topology class, then 92%, 96% and 15% of transmissions are due to a single founder variant, respectively.

We have used a combination of empirical data and phylodynamic model simulation to evaluate the role of infection stage at transmission and route of transmission on the number of virus particles transmitted during sexual HIV exposure. This makes three important advances on previous work. First, it is the first empirically-based study that fits a model to data to understand the role of the source partner in establishing infection. Second, while we use previously developed topology classification of phylogenetic trees to understand HIV transmission pairs, we extend this methodology by calibrating a phylodynamic model to empirical data. This new approach provides a means to validate the untested assumption that the number of embedded recipient partner lineages in a phylogenetic tree directly corresponds to the minimum number of founder variants transmitted. Third, our phylodynamic model explicitly incorporates virus particle number and the identity of genetic sequences. Previous work has shown that the multiplicity of founder variants has little impact on the topology class of the phylogenetic tree when only overall genetic diversity, rather than sequence identity, is tracked ^13^.

The relative importance of acute and chronic stages of HIV in determining both the number of virus particles and the number of variants transmitted is consistent with a recent modelling study ^19^. However, our study finds higher proportions of infections initiated by multiple variants overall during these two stages. This difference is likely due to the assumptions related to how the stages of infection are defined as well as the relative importance of transmission during late infection. Specifically, the previous modelling study finds that two thirds of multiple variant transmission occurs during the pre-AIDS stage of infection which is assumed to have both a high viral load and large haplotype diversity. If later stages of infection account for disproportionately less transmission then the previous model would predict higher proportions of multiple variant transmission in both the acute and chronic stages of infection, becoming more consistent with empirical estimates from our analysis. By contrast, our study is agnostic about the relative importance of early and late transmission and does not differentiate between chronic and a pre-AIDS stage of infection, which can not easily be identified through analysis of empirical data.

Data from four of the MSM transmission pairs in this study have previously been used to estimate the number of variants founding infection using a combination approach of single genome amplification (SGA), direct amplicon sequencing and mathematical modelling ^9^ This previous analysis broadly agrees with our results, with two recipients likely infected with one founder variant, one recipient with one to two founder variants and one recipient with two to three variants. Small differences likely arise because this study uses sequence data from both partners to evaluate the multiplicity of founder variants in the recipient partner. These extra data can be used to parameterize a mathematical model that accounts for the evolutionary relationship between the virus samples from both partners, rather than relying solely on accumulating diversity. Specifically, neglecting the extent of genetic similarity between the source and recipient virus samples might misattribute borderline cases of diversity accumulation.

Our study finds a median of one founder variant and a maximum of 11, with little difference between HET and MSM risk groups. When only multiple variant transmissions are considered, our study finds a median of 2-3 founder variants. These values are consistent with a previous pooled analysis using results from four analyses that used the current gold-standard SGA combination approach as above ^9^.

There are two primary limitations to acknowledge. First, our model assumes a single transmission event between each source and recipient partner. Without detailed knowledge of the transmission pairs, we cannot distinguish between multiple infections each with a single variant and a single infection with multiple variants. Second, our phylodynamic framework does not account for the effect of selection and recombination. Specifically, selection, such as that for viruses which use the CCR5 co-receptor ^20^, is thought to occur at the point of transmission, although the strength may be dependent on the route of transmission ^21^.

Our study finds that HIV-1 founder variant multiplicity is determined by infection stage of source partner, with higher founder variant multiplicity of HIV-1 in acute compared with chronic infections. These findings stress the importance of the stage of infection at transmission as an important mediating or confounding factor when assessing the role of sexual risk group in HIV epidemiology.

## Methods

### Data collation on linked transmission pairs

We automatically retrieved all HIV sequence data for men-who-have-sex-with-men (MSM) and heterosexual (HET) HIV transmission pairs for whom the direction of transmission is reported from The Los Alamos National Laboratory (LANL) HIV sequence database up to February 2019, such that each transmission pair comprise a ‘source’ and a ‘recipient’ partner (Supplementary Information). For each partner in the transmission pair we collected the following clinical and epidemiological data: (i) date of infection or time of infection prior to sampling, (ii) date of seroconversion or date of seroconversion prior to sampling, (iii) Fiebig stage at the time of sampling, (iv) date of sampling or time of sampling prior to infection, (v) number of sequences, (vi) genomic region, (vii) HIV subtype, and (viii) reported risk group. For each set of these transmission pair data we estimated, relative to the transmission time to the recipient partner (time = 0): (*i*) the time of transmission to the source partner, (*ii*) the time of the sampling of the source partner, and (*iii*) the time of sampling for the recipient partner (**Fig. 1**, Supplementary Information). We excluded all transmission pairs from further analysis for whom these three times could not be determined, for whom a risk group was not provided, for whom either partner has fewer than five sequences for all sampling times, or for whom the pair did not have a LANL cluster ID. For our base case analysis, we used the longest available genomic region with five or more sequences per partner. If more than one sampling time is available for any of the individuals, we selected the sample closest in time to the recipient transmission.

### Empirical transmission pair analysis

#### Tree reconstruction

For each of the included transmission pairs, we generated posterior sets of phylogenetic trees. For this, we first constructed alignments using Muscle v3.8.31 ^22,23^ with subtype specific reference sequences retrieved from the LANL HIV sequence database. Using these alignments, we built phylogenetic trees with MrBayes 3.2.7 ^24,25^ under the assumption of a general time-reversible (GTR) nucleotide substitution model with the addition of invariant sites (I) and a gamma distribution of site rates. We constrained sequence data to be monophyletic with respect to the reference sequences to root the tree but ingroup relationships were unconstrained to avoid any topology class bias. We ran two Markov chains each with 30 million iterations, from which we sampled every 3,000th after discarding the first 50% as burn-in which provided an average standard deviation of split tree frequencies of below 0.01 or an effective sample size of greater than 300. This gave an empirical posterior distribution of *N* = 5,000 sample trees. In a sensitivity analysis, we tested the alternative method of using maximum likelihood phylogenetic tree reconstruction with bootstrapping.

#### Empirical topology class

We classified each of the resulting phylogenetic trees in the posterior distribution as either monophyletic-monophyletic (MM), or paraphyletic-monophyletic (PM), or paraphyletic-polyphyletic (PP), to reflect the cladistic relationship between the lineages from both individuals (Supplementary Information). Each transmission pair, *k*, is then described as a triplet of probabilities, *D*_*k*_, denoting the frequency of each topology class within the *k* th pair’s posterior distribution D_k_ = {*d*_*k*_(*t*)}_*t* ∈*T*_ = {Pr (*t* | *k*)}_*t*∈*T*_ where **Σ**_*t* ∈T_ Pr(*t*|*k*)= 1 and *T* ∈ {MM, PM, PP}.

### Simulated transmission pair analysis

We simulated the transmission of virus particles and within-host evolution, accounting for the epidemiological characteristics for each transmission pair. For each transmission pair, we simulated a chain of three HIV infections: (i) an unsampled index case who infected the source after three years of their own infection during their chronic stage to reflect that the majority of both HET and MSM transmission pairs transmitted during the chronic stage (101/112 pairs). In a sensitivity analysis we accounted for the assumption that transmission rate may be higher during the acute stage, with half of the index to source transmissions occurring after 90 days and the remaining half after three years, (ii) the source individual of the transmission pair, and (iii) finally the recipient individual of the transmission pair. For each individual within each trio, we simulated viral phylogenies that reflect between- and within-host viral evolution using VirusTreeSimulator ^26^, using as input the respective epidemiological and clinical information (Supplementary Information). We used a within-host effective population size consistent with that parameterized by the PANGEA-HIV study with the following logistic model parameters: initial effective population size (*N*_0_) is 1, viral generation time (*τ*) is 1.8 days, effective population per year growth rate (*r*) is 2.85022, and time to half the carrying capacity of the viral population (*t*_50_) is 2 years ^26^. For each transmission pair, we simulated a dated viral phylogeny that has the same number of tips as the number of retrieved sequences per partner and that is sampled at the respective sampling times for the source and recipient partner (Supplementary Information). For each recipient partner infection, we assume that a total of *n*_R_ virus particles founded the infection. For each simulation, we further assume a total of *n*_S_virus particles founding infection of the source. We assume *n*_R_ takes values between one and a maximum of 12 and varied *n*_S_ between one and two (Supplementary Information). We assume that the virus samples from each recipient is representative of the within-host diversity, and that each founding virus particle has an extant lineage. Therefore, we first assigned each sample (tip) of a phylogeny as a descendant of one of the *n*_R_ virus particles. If there were more than 12 samples then the remaining tips were assigned randomly to the *n*_R_ = 12 virus particles. If there were fewer than 12 samples, then we constrained the number of founding virus particles, *n*_R_, to equal the number of samples. For every transmission pair, and for each value of *n*_R_ and *n*_S_, we simulated 100 viral phylogenies.

For every simulated viral phylogeny, we simulated transmitted sequences by adding dummy nodes with a negligibly short branch length after the transmission time. We then simulated the evolution of nucleotide sequences along the tree using Seq-Gen ^27^ and a GTR + I + gamma substitution model. The length of the simulated sequences and the evolutionary tree scaling rate match each transmission pair’s empirical sequence data. For this, we used previously estimated empirically-derived within-host evolutionary rates ^28^ and the HXB2 sequence homologous to the pair’s sequence fragment as the ancestral sequence at the root. Every transmission pair simulation produces a tip sequence alignment and a number of founder sequences equal to the number of transmitted particles.

#### Simulated topology class

We reconstructed a phylogeny using maximum likelihood inference in IQ-TREE 1.6.11 ^29^ and selected the best-fit nucleotide substitution model with ModelFinder ^30^. Each phylogeny was classified as either MM, PP or PP (Supplementary Information). Consequently, for each transmission pair *k* and each transmissibility model (i.e. number of viral particles founding infection of the recipient *n*_R_, we generated a triplet of probabilities 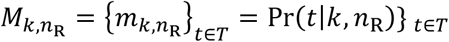 where **Σ**_*t*∈T_ Pr(*t*|*k, n*_R_)= 1 and *T* ∈ {MM, PM, PP}.

### Transmissibility model calibration

For each transmission pair, we chose the most likely value of *n*_R_ (the number of virus particles founding each recipient infection) by matching the posterior topology class from the empirical phylogenetic transmission trees with the simulated distribution of topology class. Specifically, for each transmission pair, *k*, we estimated the most likely number of viral particles founding each recipient infection 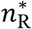 as the *n*_R_ that maximises the multinomial likelihood function 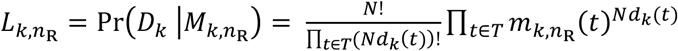. For each transmission pair *k*, we calculated lower and upper confidence limits for 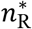 as the minimum and maximum values of *n*_R_ that satisfy 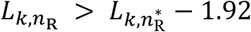 and 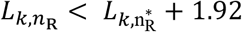, respectively ^31^. For each transmission pair *k*, we retain the best fit model for further analysis such that there are 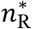 viral particles founding infection of the recipient.

### Haplotype analysis

#### Probability of a single founder haplotype

For each transmission pair, *k*, from the best fit transmissibility model, we defined the random variables 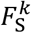 and 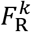 as the number of haplotypes that found infection of the source and the recipient partners, respectively. We then calculated the probability of there being a single founder haplotype in the recipient, stratified by topology class of the simulated phylogenetic tree (MM, PM, PP) and the number of founder haplotypes, *i*, in the source partner, 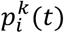, that is, 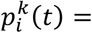. Next, we defined the probability of a single founder haplotype in the recipient as a function of a tree topology, *t*, 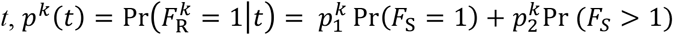. By assuming that the source partners are randomly selected from the general MSM or HET population in which the probability of a single founder strain has been calculated to be approximately 0.7 ^19^, we set, Pr(*F*_S_ = 1) =0.7 and Pr (*F*_S_ > 1) = 0.3. Finally, for each transmission pair, we calculated the probability of one founder haplotype given the observed triplet of empirical posterior topology classes *D*_*k*_, as 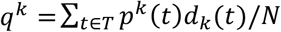.

#### Founder haplotype multiplicity by source partner infection stage

We stratified all the transmission pairs into two sets by the infection stage of the source partner. We classified the acute transmission set as those pairs for whom recipient infection is within 90 days of source infection (a set of *n*_acute_ pairs), and the chronic transmission set as those pairs for whom recipient infection is 90 days or later after source infection (a set of *n*_chronic_ pairs). For each group, we calculated the mean probability of one founder haplotype being transmitted to the recipient in each set as:

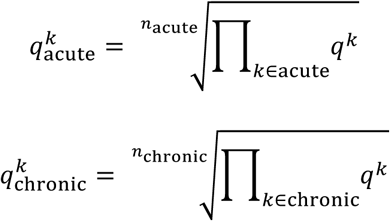

Finally, we calculated the relative risk of one founder haplotype transmitted during the acute stage versus the chronic stage by 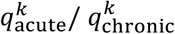.

## Data Availability

All code for data retrieval and analysis available at github.com/AtkinsGroup/TransmissionPairs_*

## Data availability

All code and data are available at github.com/AtkinsGroup under the following repositories: data on the transmission pairs and sequence alignments are provided at (TransmissionPairs_Data), code for retrieval of transmission pair epidemiological data and metadata from Los Alamos National Laboratory HIV sequence database (TransmissionPairs_LANLRetrieval), code for sequence retrieval from GenBank (TransmissionPairs_GenBankRetrieval), code for phylodynamic analysis (TransmissionPairs_PhylodynamicAnalysis), and code for topological classification (TransmissionPairs_TreeTopologyAnalysis).

## Author contributions

KEA conceived the study. CJVA, MH, KL, SH, KEA designed the study. CJVA performed the experiments and analysed the data. CJVA, MH, KL, SH, KEA interpreted the data. SGG created new software used in the study. KEA and CJVA drafted the manuscript, with critical revisions from MH, RRR, KL, SH. All authors approved the final version of the manuscript.

## Acknowledgements

CJVA and KEA were funded by an ERC Starting Grant (award number 757688) awarded to KEA. KAL was supported by The Wellcome Trust and The Royal Society grant no. 107652/Z/15/Z. MH was funded by The HIV Prevention Trials Network (grant number H5R00701.CR00.01) and The Bill and Melinda Gates Foundation (grant number OPP1175094).

## Supplementary Information for

### Methods

#### Epidemiological data and sequence retrieval

For the ease of replicating our results and using existing transmission pair data for other purposes, we developed a Python script to automatically retrieve epidemiological and metadata for each transmission pair from the Los Alamos National Laboratory HIV sequence database (LANLdb). This script downloads the following data from both the source and recipient partners to a .csv file using as input the cluster and patients ids from LANLdb: (i) time of infection, (ii) time of seroconversion, (iii) Fiebig stage at the time of sampling, (iv) number of sequences, (v) genomic region, (vi) HIV subtype, (vii) reported risk group and (viii) GenBank accession IDs.

Next we used the downloaded GenBank accession IDs to automatically retrieve (ix) viral genetic sequences and (x) sampling dates (calendar dates) from GenBank using an R script. If information from (i) to (x) were missing for any individual, we manually retrieved these values from the original manuscripts where possible.

Completed datatables from these automatic and manual processes are provided at github.com/AtkinsGroup.

#### Epidemiological data pre-processing

For each transmission pair, we define time = 0 as the time of recipient infection. We then calculated, using the data table retrieved, i) the time of infection of the source, ii) the time of sampling of the source, iii) the time of sampling of the recipient.

To estimate these times, we first calculated days from infection for both the source and the recipient partners. When these values are not given explicitly, we calculated them from time since seroconversion estimates or from Fiebig staging results. Specifically, we interpret (Cohen et al. 2010) seroconversion as the individual reaching Fiebig stage III (ELISA positive) that occurs between 22-37 days after infection (Cohen et al. 2010) and Fiebig stages I (viral RNA positive) and II (18-34) occurring 13 days and 28 days after infection, respectively. For all the pairs where a range of possible values is calculated, and for when a calendar month is provided, we incorporated the uncertainty around the infection and sampling times by assuming all values in these ranges are equally plausible and uniformly sampled within these range to account for the uncertainty.

For some pairs, the source was classified as ‘recent’ or ‘late’ at the time of transmission to the recipient partner. In these cases, we do not have an exact point to call time = 0. Therefore, for these pairs, for each simulation, we sample with replacement the time between source and recipient infections from the other pairs for whom we have previously classified as acute (<90 days delay), and chronic (90 days or more delay), sampling from the same risk group (MSM or HET) in each case.

All calculations, corresponding notes and final transmission times for each pair are provided at github.com/AtkinsGroup and visualised in **Fig. S1**.

**FIGURE S1:**
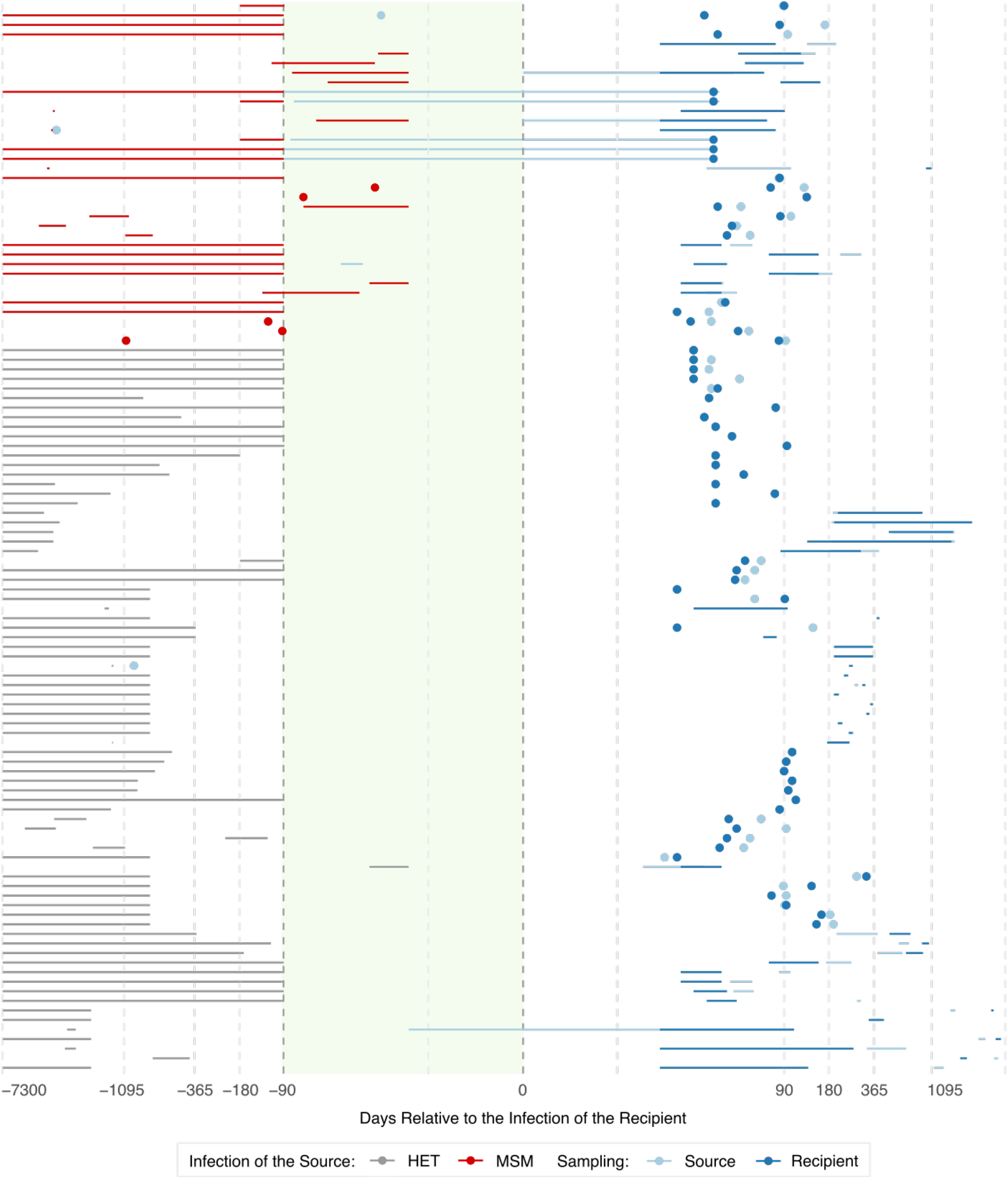
Infection and sampling times of the source and recipient for all the 112 transmission pairs analysed. Individual points denote exact times and lines denote uniform uncertainty in times. Source partners points/lines overlapping the green shaded area correspond to transmission pairs for whom transmission occurs during the acute stage.

### Results

#### Transmission pairs sequence data

Our alignments are provided at github.com/AtkinsGroup.

On average, 22 (IQR 13-33) HIV sequences are obtained from the source and 21 (IQR 10-20) sequences from the recipient for the MSM pairs, and 21 (IQR 12-25) and 18 (IQR 9-22) for the HET source and recipient, respectively. All MSM sequence data belong to subtype B, while most heterosexual sequence data belong to subtype C (49%), followed by subtype B (22%), subtype D (21%), subtype A/A-like (7%) and unclassified subtype (1%). A total of 7 (19%) of the MSM pairs have near full genomes sequenced and the remaining pairs had *env* available (mean 1653 nt, range 182-3827 nt). Ten (13%) of the HET pairs had near full genomes available, while 56 (75%) pairs had *env* (mean 1321 nt, range 323-2582 nt), nine (12%) pairs had either *pol* or *gag* (mean 1484 nt, range 1375-1499 nt) and one pair had vif-LTR3 (4666 nt) sequenced.

#### Effect of number of founding virus particles in the source

To assess whether the number of founding virus particles in the source partner affects the diversity of sequences founding infection in the recipient, we model a scenario where the index case transmitted one, two or six virus particles to the source partner at either one or three year(s) after infection. The source in turn transmits 1 to 6 virus particles to the recipient at 30 days (acute) or 1095 days (chronic) later. The simulation produces a dated viral phylogeny with tips sampled at either 30 (early) or 1065 days (late). We model 1kb nucleotide sequences along the simulated viral phylogenies using the same method as in the main text.

The genetic variation rapidly and steadily increases over time-the maximum diversity among transmitted haplotypes to the recipient was higher when the index case was infected for longer and the transmission to the recipient occurs during the chronic stage of the source (**Fig. S2**). When the source has more than one founding particle, this leads to a bimodal distribution of maximum diversity among transmitted variants within the recipient. The first and second mode represent maximum diversity when drawing the recipient founder haplotypes from either one or more than one viral population within the source, respectively. However, increasing the number of founding virus particles to more than two within the source only increases the density around the second mode without affecting the range of the maximum diversity distribution. This consistency occurs because increasing the number of founding virus particles in both transmission partners, increased the probability of drawing founding variants from different genetic pools in the source. However the average maximum diversity of the founder variants does not change because the source genetic pools evolved at the same rate and under the same evolutionary constraints with no selective advantage. This leads to genetic pools with equivalent cumulative genetic change but distinct identity. Taking this into account, we chose to model one or two founding virus particles within the source partner as we were interested in capturing some degree of variation in the transmitted haplotypes rather than multiple genetic identities *per se*.

**FIGURE S2:**
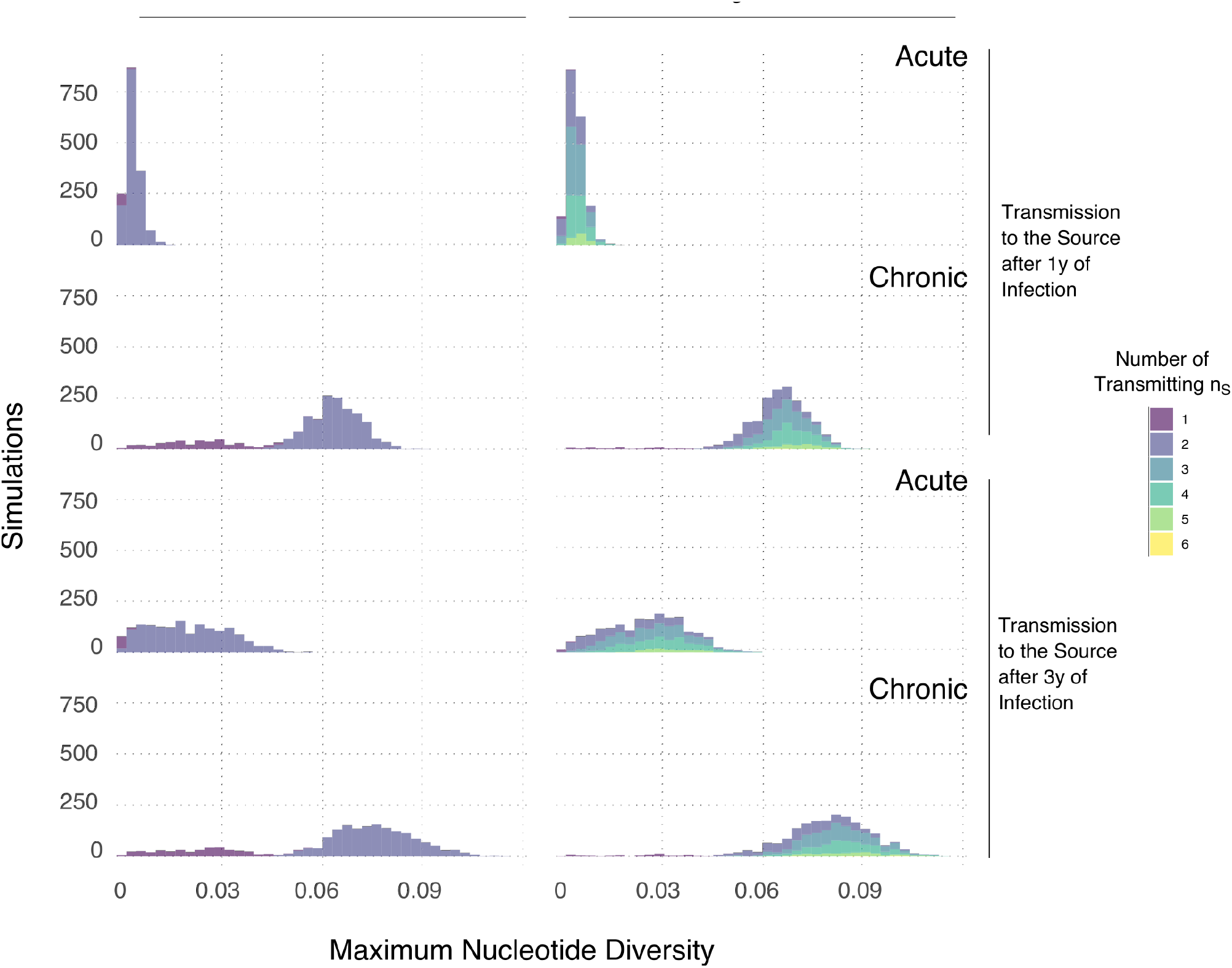
Effect of number of founding virus particles in the source.

#### Effect of using a confidence threshold for assigning the topology class

**FIGURE S3:**
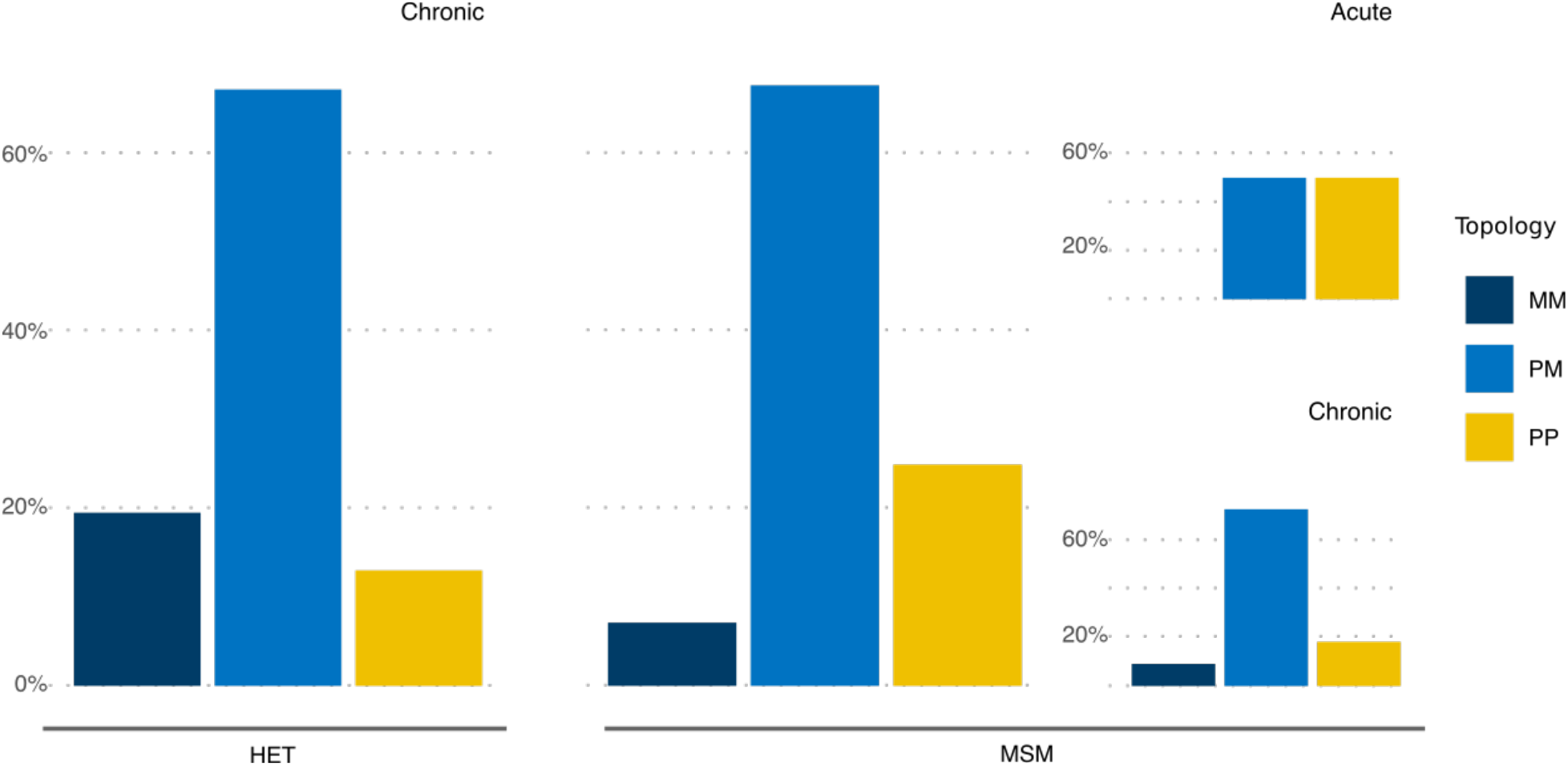
Phylogenetic findings of the empirical transmission pairs for whom the posterior trees gave a certainty of over 95% for the most frequent topology. Fraction of phylogenetic tree topology class (MM - Monophyletic-Monophyletic, PM - Paraphyletic-Monophyletic and PP - Paraphyletic-Polyphyletic) where each tree topology class is classified as the most frequent topology class of each posterior distribution per transmission pair. Results are stratified by risk group: 76 heterosexual (HET) pairs and 36 men-who-have-sex-with-men (MSM) pairs) and infection stage of the source partner at transmission (11 acute pairs defined as <90d post infection and 101 chronic pairs defined as ≥90d post infection).

#### Effect of index partner stage of infection at transmission

In the main analysis we assumed that all index cases transmit to the source partner after three years of infection. Here we also evaluated the results assuming the transmission risk was skewed towards early infection, with half of all simulations across all transmission pairs assuming index case transmission occurs during the acute stage (≤90d) and half occurs during the chronic stage (91d-3y). We find qualitatively similar results as our main analysis. The median number of variants transmitted across all pairs is 1 (range: 1-5, **Fig. S4A**). Across all pairs in both risk groups, the mean probability of observing one founder variant is 0.73. Stratifying by risk group, we find there is a higher probability that one variant founds HET infections than MSM infections (a geometric mean of 0.79 vs. 0.61, **Fig. S4B**). In contrast, when stratifying solely by infection stage of the source partner, we find that transmission during the acute stage has a much lower probability of one founder variant than during the chronic stage (means of 0.38 vs. 0.78) with a higher median number of variants transmitted, when only the most likely multiplicity for each pair is considered (2 vs. 1, **Fig. S4A**). From these results, therefore, there is still approximately twice the chance of multiple variant transmission during acute stage infection across both risk groups (relative risk is 0.48).

**FIGURE S4:**
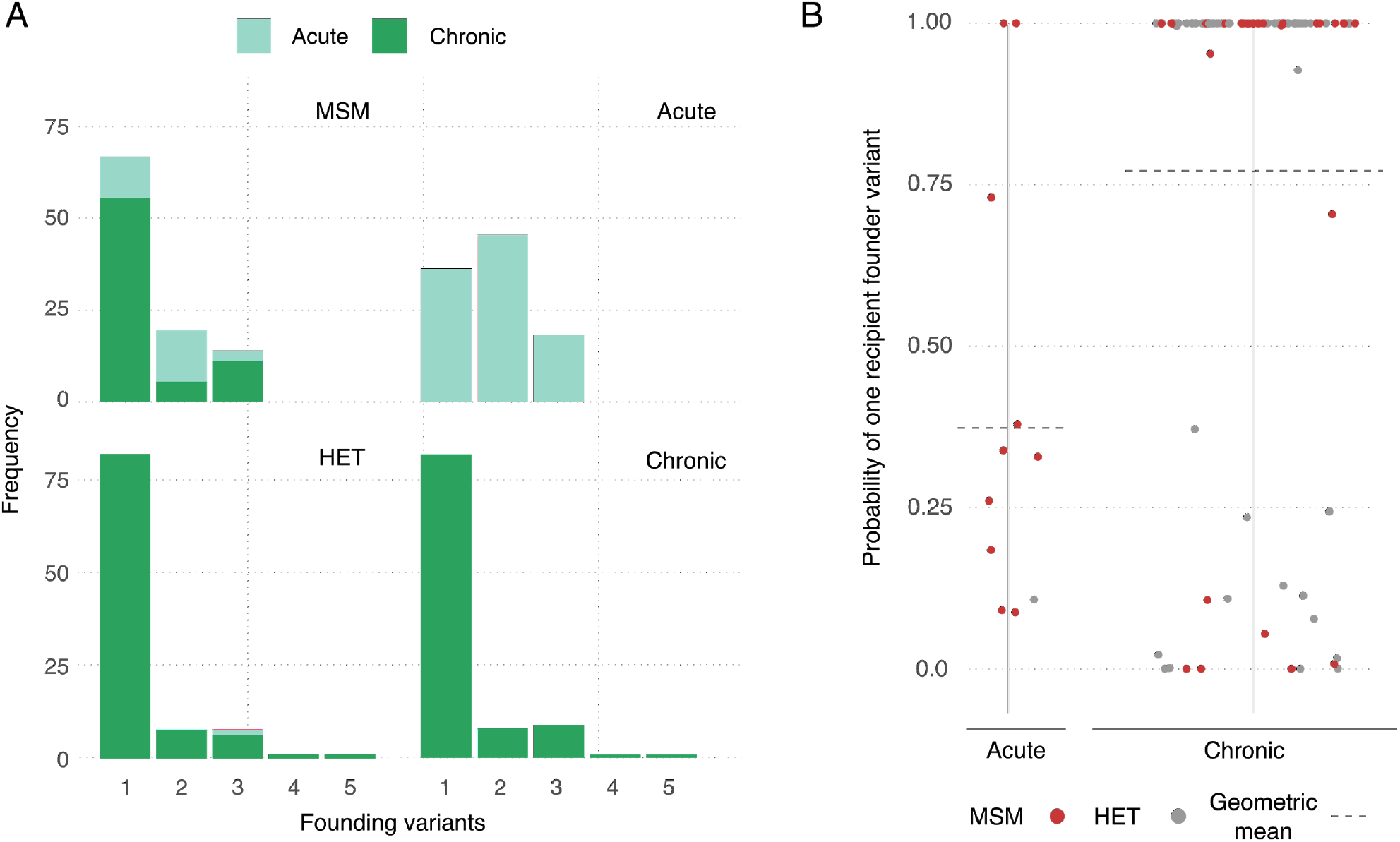
Phylogenetic findings from the calibrated simulations with skewed transmission rate towards acute stage for the index case. A) Frequency of multiplicity of founding variants for transmission pairs by infection stage of source partner at transmission and risk group. The multiplicity is calculated as the modal simulated value. B) Probability of one founder variant in the recipient for each pair stratified by infection stage of the source partner at transmission.

#### Effect of constructing empirical data phylogenetic trees using maximum likelihood inference with bootstrapping

In the main analysis we used Bayesian phylogenetic reconstruction to analyse the empirical sampled genetic data of each empirical transmission pair, using the respective posterior distribution to calculate the frequency of each topology class (MM, PM and PP) r. Here we provide a sensitivity analysis to calculate the tree topology class distribution of the empirical sampled genetic data by maximum likelihood phylogenetic tree construction and bootstrapping. After bootstrapping the empirical data 100 times to calculate the frequency of MM, PM and PP topology classes for each transmission pair, we then proceeded using the same methodology as in the main text. That is, we fit the simulation model (parameterised with the pair-specific data) to the bootstrapped data individually for each transmission pair by comparing the frequencies of tree topology classes. Overall our results remained consistent with our main results, albeit with slightly lower probabilities of observing one founder variant. The median number of variants transmitted across all pairs is 1 (range: 1-11, **Fig. S5A**). Across all pairs in both risk groups, the mean probability of observing one founder variant is 0.62. Stratifying by risk group, we find there is a higher probability that one variant founds HET infections than MSM infections (a geometric mean of 0.67 vs. 0.53, **Fig. S5B**). Stratifying by infection stage of the source partner, we find there is a much lower probability of one founder variant during the acute stage than during the chronic stage (means of 0.31 vs. 0.66) with approximately twice the chance of multiple variant transmission during acute stage infection across both risk groups (relative risk is 0.47).

**FIGURE S5:**
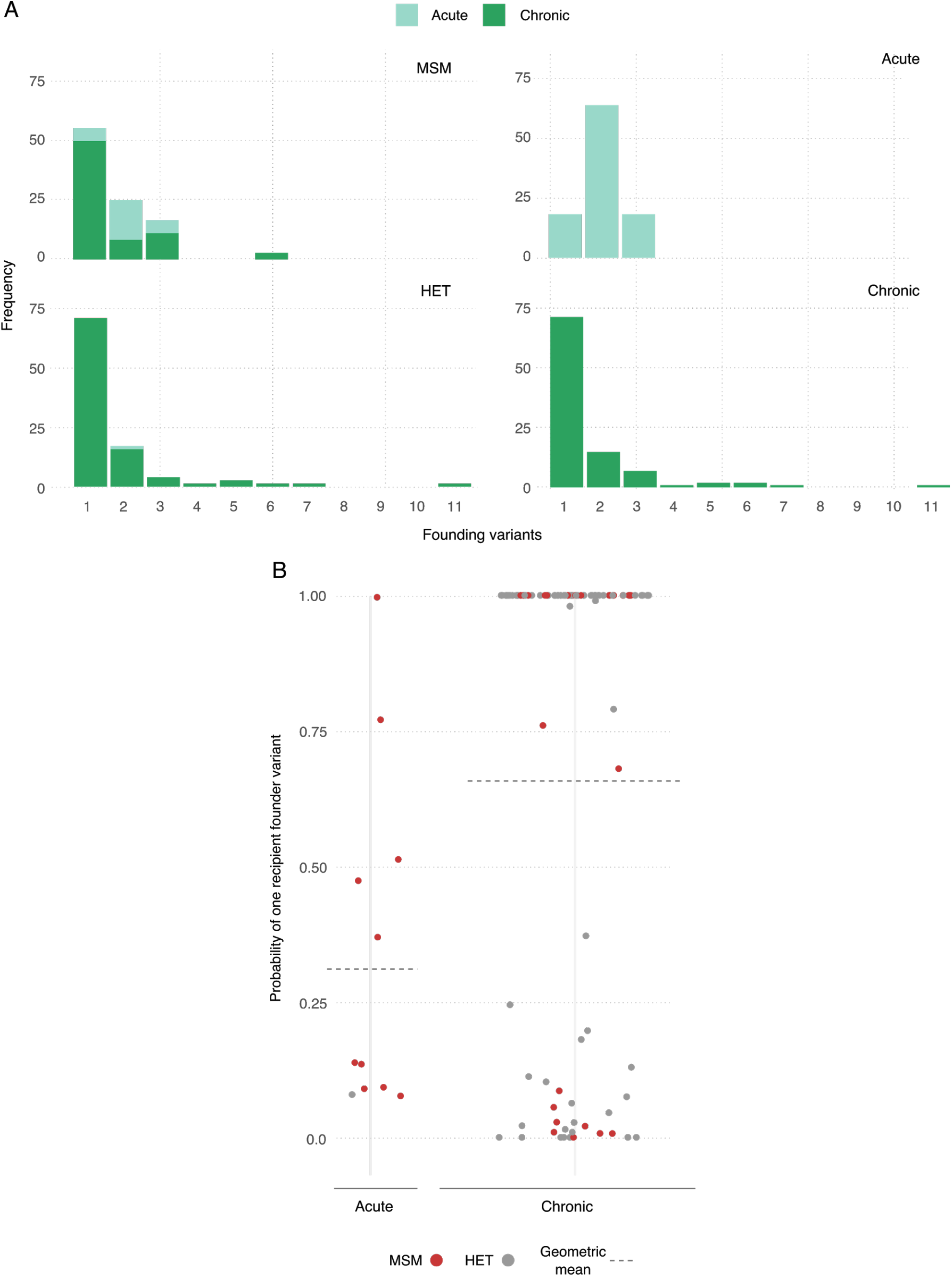
Phylogenetic findings from the calibrated simulations with bootstrapped empirical data. A) Frequency of multiplicity of founding variants for transmission pairs by infection stage of source partner at transmission and risk group. The multiplicity is calculated as the modal simulated value. B) Probability of one founder variant in the recipient for each pair stratified by infection stage of the source partner at transmission.

